# Cardiac Damage Risk in Radiotherapy of Esophageal Cancer: The Importance of Baseline Cardiac Risk Profile

**DOI:** 10.1101/2022.01.29.22269614

**Authors:** Hamid Ghaznavi, Farzaneh Allaveisi, Farzad Taghizadeh-Hesary

## Abstract

**Background:** The radiotherapy techniques are evolving. Besides optimal tumor coverage, considering organs at risk (OAR) is pertinent to radiation oncologists. In esophageal cancer radiotherapy, heart is the main OAR. Studies on excess absolute risk (EAR) of cardiovascular disease (CVD) in esophageal cancer radiotherapy are limited in the main literature. Therefore, this study was conducted to estimate the EAR of CVD in patients with esophageal cancer treated with the IMRT technique.

**Materials and Methods:** Seventeen patients with esophageal cancer were selected. The patients were planned for the IMRT technique, and the 10-year EAR of CVD was estimated using the linear model. The data of major CVD risk factors [including age, smoking, and family history of heart attack, systolic blood pressure, total and HDL cholesterol, and high sensitivity c-reactive protein (hsCRP)] were obtained and the baseline risk of CVD was categorized into low- and high-risk groups using the Reynolds risk score.

**Results:** Family history of heart disease and smoking increased the EAR of CVD significantly compared to the cholesterol and hsCRP. The 10-year EAR of the high-risk group was more than four times of the low-risk group at all ages. In the low-risk group, EAR of CVD after radiotherapy of esophageal cancer can increase by up to 9.1%, while in the high-risk group, EAR increased by 34.89%.

**Conclusions:** Adding the baseline CVD risk factors improved the estimation of EAR of heart disease after esophageal cancer radiotherapy with the IMRT technique.

## 1. Introduction

Chemoradiotherapy (CRT) is a standard of care for patients with esophageal cancer (EC) in both definitive and pre-operative settings. Intensity-modulated radiation therapy (IMRT) and 3-dimensional conformal radiation therapy (3D-CRT) are two standard radiotherapy techniques for EC ^[1]^. In comparison, IMRT allows selective tumor targeting and sparing of organs at risk (OAR), such as heart ^[2, 3]^. Considering the heart’s proximity to the esophagus, the risk of CVD should be considered. The incidence of cardiac toxicity mainly depends on the target volume and the tumor location, as well as the irradiation technique. This might be in the form of pericardial disease, cardiomyopathy, coronary artery disease, valvular heart disease, and arrhythmias. The literature shows that the incidence of cardiac complications in this setting varies widely (range: 5-44%, mean: 10.8%), which mainly occurs within the first three years of irradiation ^[4-6]^. In addition to the absorbed radiation dose, the development of heart disease depends on CVD risk factors such as diabetes mellitus, smoking, family history of heart disease, and several other factors, such as systolic blood pressure, total and high-density lipoprotein (HDL) cholesterol, and high sensitivity C-reactive protein (hsCRP) ^[7, 8]^.

A comparative study between IMRT, helical tomotherapy (HT), and volumetric modulated arc therapy (VMAT) techniques noted that IMRT is superior in terms of normal tissue complication probability (NTCP) of heart in middle thoracic EC, HT had higher dose conformity and homogeneity in cervical EC, and VMAT reduced the liver and lungs NTCPs in lower thoracic EC ^[9]^. Another study aimed to develop and validate NTCP models for cardiac and pulmonary toxicities as well as one-year mortality after preoperative CRT in EC patients. The results confirmed the validity of developed models for pulmonary toxicity and mortality, but accurate prediction of cardiac complications was not obtained ^[10]^.

The importance of developing radiobiological models in radiotherapy is to estimate the NTCP and guide selecting the optimum technique ^[11]^. Studies have well established that the baseline CVD risk factors have a determining role in developing cardiac toxicities ^[7, 12]^. However, there is no study in the literature incorporating the baseline CVD risk factors to estimate the probability of radiation-induced cardiac toxicity in EC patients to the best of our knowledge. This study was therefore designed to calculate the excess absolute risk (EAR) of CVD after radiotherapy of EC with considering the baseline risk of CVD.

## 2. Materials and Methods

### 2.1. Treatment planning

Computed tomography (CT) data sets of 17 male EC patients who received preoperative CRT were selected for treatment planning. The patients were scanned with 3□mm slice thickness in a simulation CT scan (GE LightSpeed CT simulator). The CT datasets were transferred to the ISOgray version 4.1.3.23L treatment planning system. Patients were treated on the Elekta Synergy® accelerator using 6-MV photons. The target and heart volumes were delineated according to the Radiotherapy and Oncology Group (RTOG) guideline ^[13]^.

For each patient, the field-in-field forward planned (FIF-FP) IMRT technique—including four radiation fields (anterior, posterior, and two lateral fields)—was created. After ensuring full coverage of planning target volume (PTV), the heart dose was kept at the minimum value. Dose-volume histograms (DVHs) were obtained for the PTV and heart volume. The prescribed dose for all recruited patients was 45.0 Gy in 25 fractions.

### 2.2. Cardiovascular disease risk assessment

To estimate the excess absolute risk (EAR) of CVD (Including myocardial infarction, coronary revascularization, or death from ischemic heart disease) after radiation therapy, the following formula was applied:

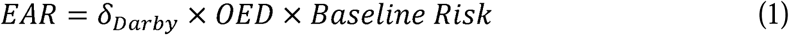

The coefficient *δ*_*Darby*_ = 0.074 *Gy*^−1^ shows the linearity of the rate of heart toxicity with increasing mean heart dose, which may occur from the first year after radiotherapy to at least the following 20 years ^[14]^. Since OARs receive a non-uniform dose, we required to convert it to a uniform dose distribution throughout the entire volume of OARs. To this end, a parameter called organ equivalent dose (OED) was used, which was considered equal to the mean heart dose and was obtained using the following formula ^[15]^:

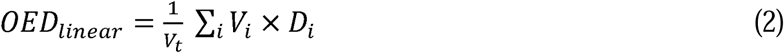

Where *V*_*t*_ is the total volume of the organ of interest and *V*_*i*_ is the volume of bin dose of the organ exposed to a homogeneous dose of *D*_*i*_. The linear model of OED assumes a linear correlation of radiation-induced CVD over the whole dose range.

The baseline CVD risk was calculated using the Reynolds Risk Score (RRS). The RRS is based on age, sex, systolic blood pressure, total and HDL cholesterol, smoking status, hsCRP level, and parental history of a heart attack before the age of 60 years. The original study of RRS was conducted in females ^[7]^. Further studies revealed that the same scoring can be implemented for male patients ^[16]^. The RRS was calculated for each subject by the score sheet available as an online calculator at: http://www.reynoldsriskscore.org/

According to Reynolds’ model, 10-year cardiovascular disease risk (%) is calculated by the following formula:

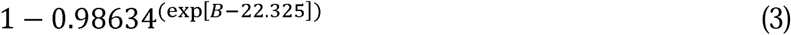

where B = 0.0799 × age + 3.137 × natural logarithm (systolic blood pressure) + 0.180 × natural logarithm (high-sensitivity C-reactive protein) + 1.382 × natural logarithm (total cholesterol) − 1.172 × natural logarithm (high-density lipoprotein cholesterol) + 0.134 × hemoglobin A1C (%) (if diabetic) + 0.818 (if current smoker) + 0.438 (if family history of premature myocardial infarction).

In this study, patients without diabetes mellitus were included to follow the RRS’s eligibility criteria. The RRS was categorized into low- and high-risk groups based on the values demonstrated in Table 1. Then, we categorized the RRS for ages 40, 50, 60, and 70 years.

**Table 1.** Risk stratification for cardiovascular disease (based on Reynolds risk score)

|  | SPB<br>(mmHg)<br>(median) | Total<br>Cholesterol<br>(mg/dL)<br>(median) | HDL<br>cholesterol<br>(mg/dL)<br>(median) | CRP<br>(mg/dL)<br>(median) | Positive<br>family<br>history<sup>a</sup><br>(yes/no) | Smoking<br>(yes/no) |
| --- | --- | --- | --- | --- | --- | --- |
| Low-risk group | 125 | 208 | 62 | 2 | No | No |
| High-risk group | 135 | 235 | 43 | 4.3 | Yes | Yes |

Abbreviations: CRP, C-reactive protein; HDL, high-density lipoprotein; SBP, systolic blood pressure.
<sup>a</sup> Parental history of heart attack before age 60

### 2.3. Ethics statement

The ethical approval was provided by the ethical committee of the Kurdistan University of Medical Sciences, and the study was conducted per the principles of the Declaration of Helsinki and current ethical guidelines.

## 3. Results

Based on the DVH analysis, the mean (± SD) total dose of PTV and heart were 45.54 (± 0.68) Gy and 15.10 (± 3.90) Gy, respectively. Overall, the mean OED of heart was 10.25 ± 2.91. Figure 1 shows dose distribution in all views of the FIF-FP plan.

**Figure 1.**
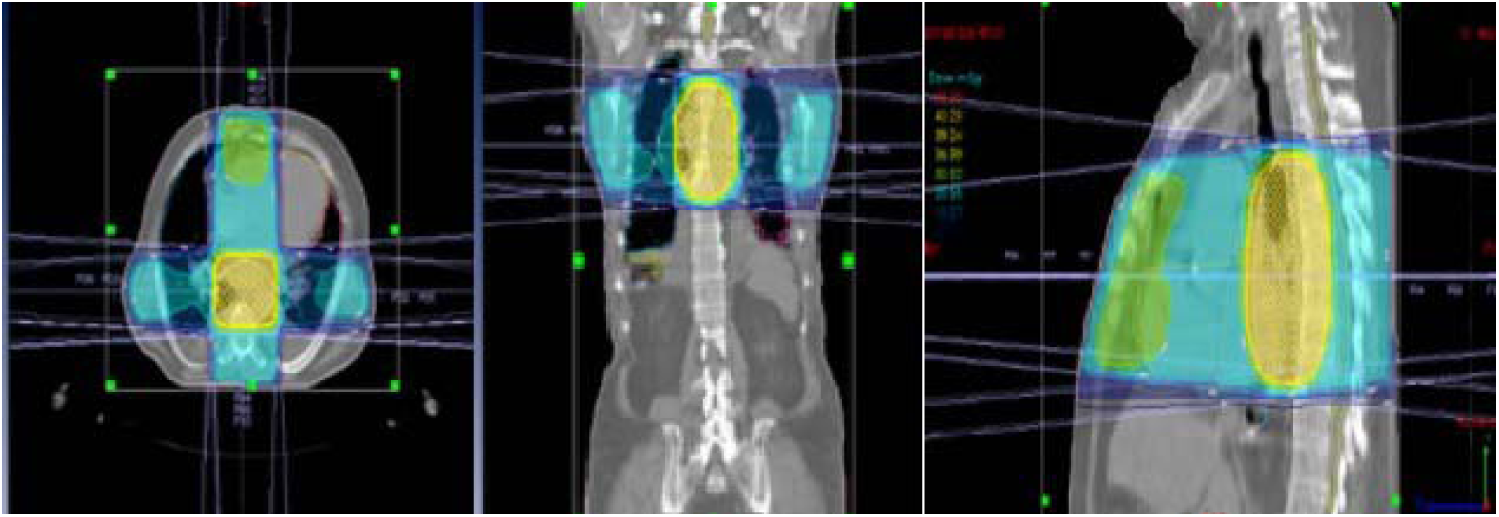
Dose distribution in three views of a patient with mid-thoracic esophageal cancer (field-in-field forward planned IMRT technique).

The calculated 10-year baseline risk for CVD for different age groups in low- and high-risk groups are summarized in Table 2. At younger ages, the effects of smoking and a family history of heart disease were almost identical. As the age increased, the impact of these factors on increasing baseline risk and consequently EAR was clearer compared to patients whom have not had these factors.

**Table 2.** The 10-year baseline risk for ages 40, 50, 60, and 70 years with considering the family history of cardiovascular disease and smoking in the high-risk group.

| Age group,<br>years | 40 | 50 | 60 | 70 |
| --- | --- | --- | --- | --- |
| Low-risk | 2% | 3% | 6% | 12% |
| High-risk | 9% | 13% | 27% | 46% |

The 10-year EAR for CVD for different age groups in low- and high-risk groups are summarized in Figure 2. In the low-risk group, EAR of heart disease after EC radiotherapy can increase by up to 9.1%, while in the high-risk group, EAR increased by approximately 34.89%. As evident, the difference between low- and high-risk groups for 10-year EAR increases with age. In all age groups, the EAR of the high-risk group was approximately more than four times of the low-risk group.

**Figure 2.**
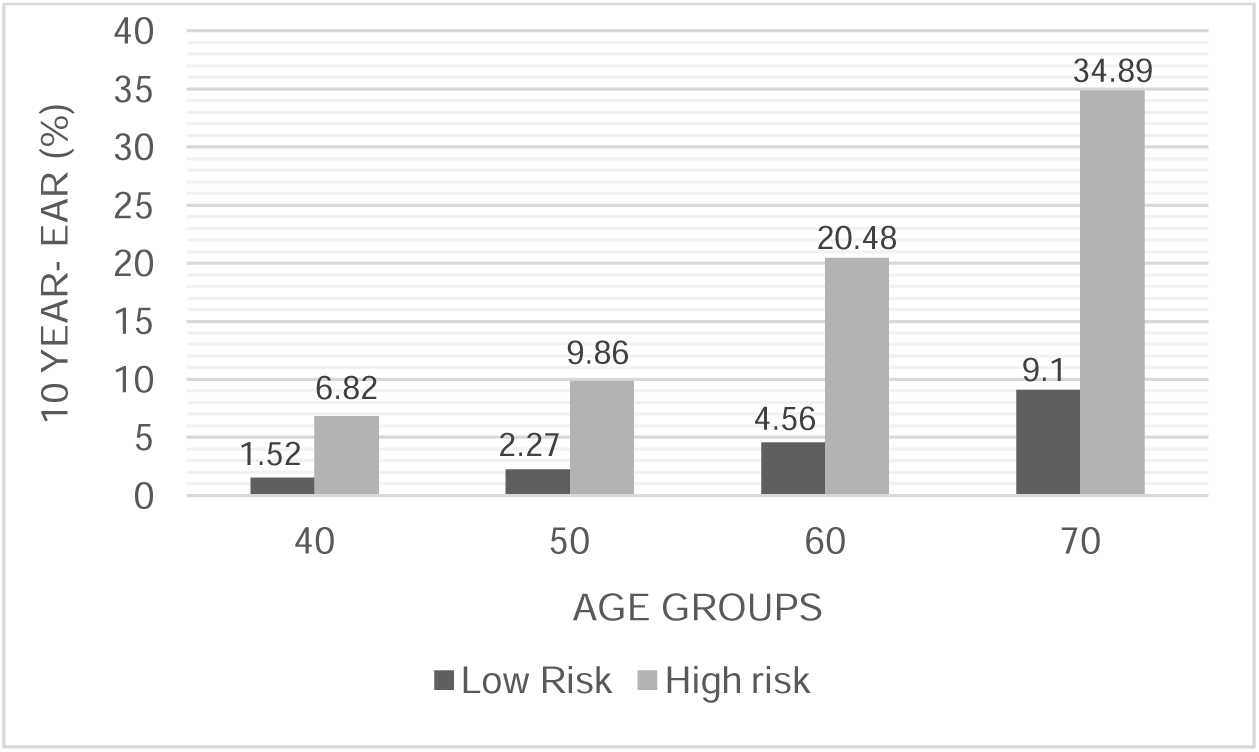
The 10-year excess absolute risk (EAR) of cardiovascular disease per the Reynolds risk groups for patients with esophageal cancer receiving radiotherapy with IMRT technique.

## 4. Discussion

The current study assessed the risk of CVD in EC radiotherapy, focusing on the baseline risk of heart diseases. We applied the IMRT technique due to its superiority to 3D-CRTin terms of target volume coverage, conformity index, and homogeneous dose distribution ^[2, 11]^. In addition, the received dose of OAR such as heart and lungs is lower in IMRT compared to 3D-CRT. These benefits have resulted in improved survival rates of EC patients receiving radiotherapy with the IMRT technique ^[17, 18]^. In this study, we found that the increased EAR of cardiac toxicities is mainly due to smoking and a significant family history of a heart attack. Considering the CVD risk factors can guide the clinicians to minimize the radiation to the heart where the target volume is adjacent to the heart.

Several studies have applied a similar approach to breast cancer. One study evaluated the 10-year EAR of heart disease in patients with breast cancer per the CVD risk profile. It showed that the 10-year EAR of CVD in medium-and high-risk groups is 0.5% and 6.0% in 50 years old, and 2.8% and 26% at 70 years old, respectively. ^[19]^. This finding reflects the importance of baseline risk of CVD in predicting the EAR of heart disease. Another experimental study demonstrated the importance of baseline CVD risk in estimating the 10-year EAR of cardiac toxicity in patients with left-sided breast cancer. The authors found that diabetes mellitus and smoking are two major risk factors of radiotherapy-induced heart damage ^[20]^. In our study, positive family history and smoking were the major risk factors of 10-year CVD.

In 2017, Allaveisi et al. demonstrated that the NTCP for cardiac toxicity in EC radiotherapy (using FIF technique) was less than 0.01% ^[11]^. However, this study ignored the effect of baseline cardiac risk profile on radiation-induced cardiac toxicity. In the current study, we considered the baseline cardiac risk—using the Reynolds model— and found that the 10-year EAR of CVD varies between 2-46% based on the cardiac risk profile.

To the best of our knowledge, a few studies have estimated the risk of heart toxicity by a radiobiological model in EC. The strength of the present study was considering baseline CVD risk to calculate the risk of cardiac damage in radiotherapy. The limitations of this study are the small sample size, lack of access to an advanced therapeutic technique such as VMAT, and not examining the effect of flattening filter-free (FFF) mode and deep inspiration breath-hold.

## 5. Conclusions

We showed the importance of considering the baseline cardiac risk profile in estimating the probability of radiation-induced cardiac damage in patients with EC. We also showed that family history of heart disease and smoking are the major determinants of EAR of CVD in this setting. Adding the cardiac risk factors significantly increased the 10-year EAR of CVD. Therefore, the baseline cardiac risk profile should be incorporated into daily clinical practice.

## Data Availability

All data produced in the present study are available upon reasonable request to the authors

## Declarations

### Availability of data and material

Not applicable

## Acknowledgement

None

